# Moving beyond risk ratios in sibling analysis: estimating clinically useful measures from family-based analysis

**DOI:** 10.1101/2025.05.16.25327702

**Authors:** Viktor H. Ahlqvist, Hugo Sjöqvist, Arvid Sjölander, Daniel Berglind, Paul C. Lambert, Brian K. Lee, Paul Madley-Dowd

**Author notes:** Corresponding author: Viktor H. Ahlqvist.

## Abstract

**Objective:** Findings from family-based analyses, such as sibling comparisons, are often reported using only odds ratios or hazard ratios. We demonstrate how this can be improved upon by applying the marginalized between-within framework.

**Study Design and Setting:** We provide an overview of sibling comparison methods and the marginalized between-within framework, which enables estimation of absolute risks and clinically relevant metrics while accounting for shared familial confounding. We illustrate the approach using Swedish registry data to examine the association between maternal smoking and infant mortality, estimating absolute risk differences, average treatment effects, attributable fractions, and numbers needed to harm (or treat).

**Results:** The marginalized between-within model decomposes effects into within-and between-family components while applying a global baseline across all families. Although it typically yields similar relative estimates to conditional logistic or stratified Cox regression, the model’s specification of a baseline enables the estimation of absolute measures. In the applied example, absolute measures provided more interpretable and policy-relevant insights than relative estimates alone. Code for implementation in Stata and R is provided.

**Conclusion:** The marginalized between-within framework may strengthen the interpretability of family-based analysis by enabling absolute and policy-relevant estimates for both binary and time-to-event outcomes, moving beyond the limitations of solely relying on relative effect measures.

**What is new?:** *Key Findings:*

- Findings from sibling analyses are typically presented using only relative measures, such as odds ratios or hazard ratios, limiting interpretability.
- This study illustrates how the marginalized between-within framework can be used to derive clinically relevant absolute effect measures while adjusting for shared familial confounding.

*What this adds to what was known?:*

- Unlike conventional methods, this approach enables estimation of absolute risks, average treatment effects, attributable fractions, and numbers needed to treat or harm—using standard software—while accounting for unmeasured familial confounding.

*What is the implication and what should change now?:*

- Researchers conducting sibling comparisons should consider adopting the marginalized between-within framework to report both relative and absolute effect measures.
- This shift could enhance the clinical and public health relevance of family-based designs by improving interpretability and communication of findings.

## Introduction

Family-based designs, such as sibling comparisons, are powerful tools to control for unmeasured confounding shared within families.^1^ Typically, when the outcome is binary, sibling analyses are conducted using conditional logistic regression; for time-to-event outcomes, stratified Cox regression is commonly employed. However, most studies using these approaches report only hazard or odds ratios that are conditional on shared familial factors.

These ratio measures, while statistically valid, are often clinically uninformative, as they do not reflect the underlying absolute risk. For instance, consider two populations each followed up for one year. In population A, 6% of the exposed and 3% of the unexposed experience the outcome. In population B, 15% of the exposed and 8% of the unexposed experience the outcome. Despite these stark differences in absolute risk, the odds ratios are nearly identical (OR_A_ = 2.06; OR_B_ = 2.03).

One solution is to estimate absolute measures, such as marginal survival functions adjusted for confounders, which can be plotted over time—akin to Kaplan–Meier curves in randomized trials, but with adjustment for confounders in observational studies. While the theoretical foundations for such estimation exist^2,3^, these methods are rarely applied to either binary or time-to-event outcomes^4–7^. The reason why such clinically valuable measures are not calculated in sibling analysis of time-to-event or binary outcomes is not clear, but may relate to a lack of accessible methodological guidance.^4^

Here, we describe how to estimate absolute measures using a marginal between-within framework that accounts for shared familial confounding. We demonstrate how clinically meaningful quantities—such as the average treatment effect, attributable fraction, and number needed to treat—can be estimated while adjusting for both observed non-shared and unobserved shared confounding. To support applied researchers, we provide reproducible Stata and R code for both binary and time-to-event outcomes.

## Methods

In the following sections, we first outline the traditional models used in sibling analysis: conditional logistic regression for binary outcomes and stratified Cox regression for time-to-event outcomes. We then describe the between-within framework and the marginalization of such models, showcasing how one can reformulate conditional logistic regression and stratified Cox regression using the marginalized between-within framework. This reformulation eases the estimation of a baseline function, which in turn allows for the calculation of absolute risk and other clinically meaningful measures.

### Traditional models for sibling analysis

The most widely used model for binary outcomes in epidemiology is logistic regression, which, in the context of sibling analysis, is typically implemented as conditional logistic regression:

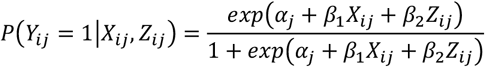

Here, *P*(*Y*_*ij*_ = 1|*X*_*ij*_, *Z*_*ij*_) is the probability that individual *j* in family *i* experiences the outcome conditional on their exposure *X*_*ij*_ and a non-shared confounder *Z*_*ij*_; *α*_*i*_ is a family-specific intercept which absorbs all shared familial confounding; and *exp*(*β*_1_) represents the conditional odds ratio associated with a one-unit increase in the exposure. Although conditional logistic regression is straightforward to implement in most software (e.g., *clogit* in Stata), a key limitation is that the cluster-specific intercept *α*_*i*_ is not estimated directly. As a result, it is not possible to recover the baseline probability of the outcome, making it difficult to compute absolute risk or derived clinical measures, such as the number needed to treat.

The most widely used survival model in epidemiology is the Cox proportional hazards model.^8^ In sibling analyses, this model is typically implemented as a stratified Cox regression:

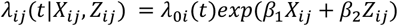

where *λ*_*ij*_(*t*|*X*_*ij*_, Z_ij_) is the individual-specific hazard function, and *λ*_0*i*_(*t*) is a family-specific baseline hazard, which absorbs unmeasured confounding shared between siblings. The corresponding survival function is given by: *S*_*ij*_(*t*) = exp (−Λ_*ij*_(*t*)), where 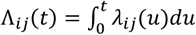. As with the conditional logistic model, the baseline hazard is not directly estimated when fitting a Cox model. Consequently, only relative effects—such as the hazard ratio *exp* (*β*_1_), which reflects the association between exposure and outcome conditional on shared family factors—are typically reported. This lack of a directly estimated baseline complicates the derivation of absolute risk measures, such as survival probabilities or risk differences, which are often of greater clinical relevance than the hazard ratio.

### The Between-Within framework

Unlike conditional logistic regression and stratified Cox regression, the between-within framework explicitly decomposes the total exposure–outcome association into a within-family (causal) effect and a between-family (shared confounding) effect. The conditional between-within model can be expressed as:

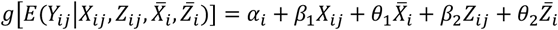

Here, 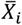 and 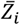 denote the family means of the exposure and non-shared confounders, respectively. The family-specific intercept *α*_*i*_ is assumed to follow a normal distribution with constant mean μ and variance σ^2^. The function *g*(·) represents a link function appropriate to the outcome (e.g., logit). Under this model, *β*_1_ captures the within-family effect of the exposure—interpreted as the association between the outcome and a 1-unit increase in exposure, controlling for observed non-shared confounders (*Z*) and all family-shared factors. This is often referred to as the within-effect. In contrast, 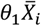 and 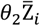 absorb the family-level confounding, which is sometimes referred to as the between-effect(s). However, estimating the random intercept *α*_*i*_ (sometimes referred to as shared frailty) can be computationally intensive and may result in unstable estimates.^1,3^ As a practical alternative, one can instead use the marginal between-within model,^9^ specified as:

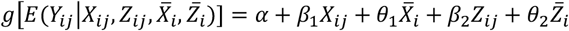

In this specification, *α* is a fixed intercept common to all families. The marginal between-within model can be viewed as an approximation to the conditional version,^3^ although they are not strictly the same. One important difference between them is that the marginal between-within model also circumvents the assumption off no effect measure modification by shared confounders.^9^ However, the marginal between-within model will be similar to the conditional between-within model if the variance in *α*_*i*_ is small^9^ and there is no selection bias due to discordance.^3^

### Marginal between-within logistic regression

Instead of using conditional logistic regression for binary sibling analysis, one can specify a marginal between-within logistic model as:

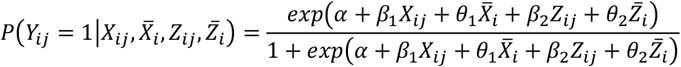

Here, *α* is a fixed intercept common to all families. The coefficients *β*_1_ and *β*_2_ capture within-family effects, while *θ*_1_ and *θ*_2_ account for between-family effects. Although the method is referred to as the *marginal* between-within approach, the term “marginal” refers to the marginalization over families, not the effect estimates themselves, which remain conditional odds ratios. Nevertheless, a practical advantage of this approach lies in its implementation: by including both within-and between-family terms, a standard logistic regression model becomes a sibling model. Furthermore, the use of a global intercept reduces the computational complexity greatly and eases the estimation of clinically relevant absolute measures. For example, from this model it is possible to estimate the outcome risk under hypothetical scenarios—such as if all individuals were exposed versus if all were unexposed—while adjusting for both observed non-shared confounders and unmeasured shared familial factors (i.e., via standardization or the parametric g-formula with control for shared familial confounding). Assuming no other confounding beyond these factors, the contrast between the standardized risks represents the average treatment effect. Moreover, one can estimate the population attributable fraction (i.e., the proportion of cases in the population attributable to the exposure) and the number needed to harm (i.e., the number of ‘treatments’ required to cause one additional outcome in the population as a whole), all while accounting for shared family-level confounders (Appendix). Of course, these are only some of the large numbers of possible absolute clinical measures one could calculate given that a baseline risk has been estimated (e.g., the average treatment effect among the factually treated, number needed to treat among the factually untreated, the attributable fraction among the factually treated, etc.)^10^.

### Marginal between-within Cox regression

The main strength of stratified Cox regression is that it does not require direct estimation of the baseline hazard. However, this also becomes a limitation when one is interested in absolute risks or survival probabilities, as the baseline hazard is necessary to compute such measures. One alternative is to introduce a cluster-specific frailty term and assume a parametric form for the baseline hazard (as Dahlqwist et al. ^4^ do for the conditional between-within model). However, this approach can be computationally intensive and potentially unstable. Instead, we can define a marginal between-within Cox model:

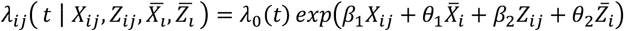

Here, *λ*_0_(*t*) is a shared baseline hazard across all families. The coefficients *β*_1_ and *β*_2_ represent within-family effects, while *θ*_1_ and *θ*_2_ capture between-family effects (note: as with the logistic regression, these are conditional log(hazard ratios) even if the model is referred to as “marginal”). Rather than estimating a separate frailty term per family^4^, we can use Breslow’s estimator^11^ to derive a cumulative baseline hazard function: 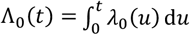. Alternatively, one may flexibly model *λ*_0_ (*t*) or Λ_0_(*t*) using restricted cubic splines (see Appendix).

Regardless of how we go about estimating the baseline common to all families, the advantages of the marginal between-within Cox model compared to the stratified Cox regression become especially clear when estimating clinically relevant measures. With a common baseline hazard estimated across all families, we can standardize survival functions to compare counterfactual risks through time — e.g., estimating the cumulative incidence of the outcome at time *t* under a scenario where everyone is exposed compared to a scenario where no one is exposed, adjusting for both observed non-shared and shared familial confounding (i.e., G-computation based on a sibling model). The contrast between these corresponds to the average treatment effect by time *t*, assuming no other confounders. In addition, access to the baseline hazard enables estimation of policy-relevant quantities such as the population attributable fraction and number needed to treat or harm, either at specific time points (e.g., 10 years post baseline) or over the entire follow-up period (see Appendix).

### Applied example: Maternal Smoking and Infant Mortality

To illustrate the utility of the marginal between-within framework, we examined the association between maternal smoking during pregnancy and infant mortality. Although the harms of maternal smoking are well established, confounding shared between siblings— such as maternal health and socioeconomic status—may influence both smoking behaviors and infant mortality risk. Sibling analysis offers a way to adjust for these shared familial confounders. Because non-twin siblings differ in maternal age, and because maternal age may affect both smoking behavior and infant outcomes, we additionally adjust for maternal age as a non-shared confounder. For pedagogical purposes, we analyzed this example using both binary and time-to-event outcome models. Specifically, we applied the marginal between-within analogs of logistic regression and Cox regression, which allow us to estimate both relative and absolute measures of association. These included the odds ratio, hazard ratio, absolute risk difference, attributable fraction, and number needed to treat. For comparison, we also fit the conventional sibling models—conditional logistic regression and stratified Cox regression—albeit these only estimate relative measures (i.e., odds and hazard ratios). To account for within-family correlation, we used cluster-robust standard errors. We estimated standard errors for absolute measures using the delta method.

We identified all live-born children in the Swedish Medical Birth Register between 1980 and 2020 with maternal smoking status recorded at the first antenatal visit (self-reported). From this cohort, we selected 2,818,660 children who had at least one full sibling born during the same period, yielding a total of 1,214,946 families. These individuals were linked to the Swedish Cause of Death Register to ascertain any recorded death. Follow-up was defined as the period from birth until one year of age, death, or December 31, 2021, whichever came first. Between 1980 and 2020, we observed 9,565 infant deaths (0.34%). Of the 2,818,660 children, 3,225 were exposure and outcome discordant.

### Example of result description based on time-to-event analysis

The odds ratios and hazard ratios were similar between the conventional sibling models and the marginal between-within models (Table 1). When using the marginal between-within Cox regression, which adjusts for all unmeasured factors shared between siblings and the observed non-shared covariate of maternal age, we found that maternal smoking during pregnancy was associated with an increased rate of infant mortality. The estimated hazard ratio was 1.68 (95% CI: 1.46–1.93) (Table 1). In terms of absolute risk, we observed a mortality difference of 0.21 percentage points (95% CI: 0.14–0.28) by day 365 of follow-up (Figure 1, Table 1). This corresponds to an attributable fraction of 8.62% (95% CI: 6.68–10.56), and a number needed to harm of 476 (95% CI: 320–632).

**Table 1.** Association between maternal smoking and infant mortality^a^ estimated using conventional (conditional logistic and stratified Cox) and marginal between-within (Cox and logistic) sibling-analysis.

|  | **Binary analysis** |  | **Survival analysis** |  |
| --- | --- | --- | --- | --- |
|  | **Conditional logistic** | **Marginal between-within logistic** | **Stratified Cox** | **Marginal between-within Cox** |
| **Conditional odds ratio, (95% CI)** | 1.63 (1.42-1.87) | 1.68 (1.46-1.94) | n/a | n/a |
| **Conditional hazard ratio, (95% CI)** | n/a | n/a | 1.60 (1.40-1.83) | 1.68 (1.46-1.93) |
| **Risk among unexposed, % (95% CI)** | - | 0.31 (0.30-0.32) | - | 0.31 (0.30-0.32) |
| **Risk among exposed, % (95% CI)** | - | 0.52 (0.46-0.58) | - | 0.52 (0.46-0.58) |
| **Risk difference, % (95% CI)** | - | 0.21 (0.14-0.28) | - | 0.21 (0.14-0.28) |
| **Attributable fraction, % (95% CI)** | - | 8.62 (6.68-10.56) | - | 8.62 (6.68-10.56) |
| **Number Needed to Harm, N (95% CI)** | - | 476 (320-631) | - | 476 (320-632) |
| <sup>a</sup>All absolute measures were calculated up until the 365<sup>th</sup> day of life. |  |  |  |  |

**Figure 1.**
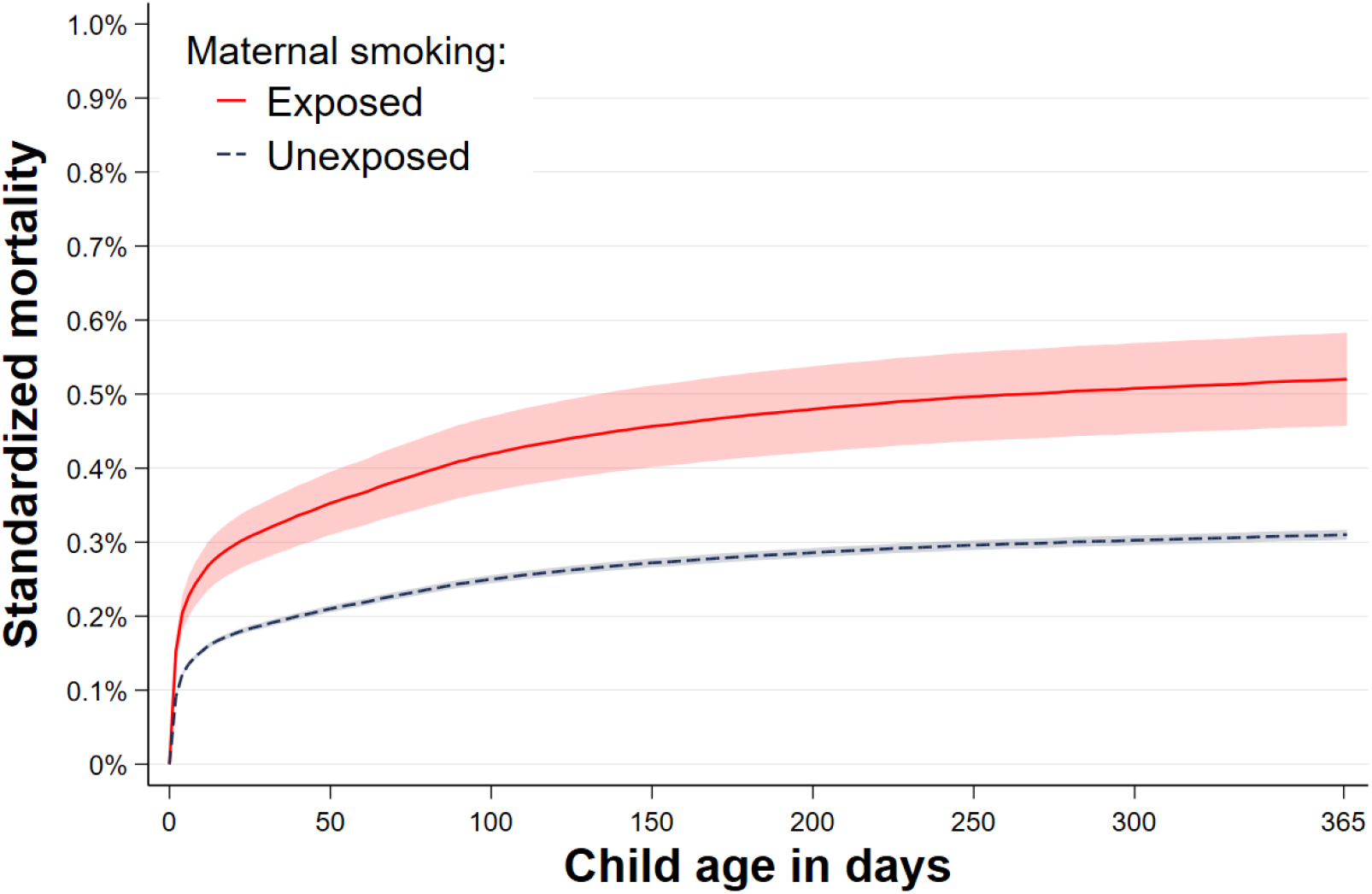
The standardized child mortality across child age according to exposure to maternal smoking, as obtained from the marginal between-within Cox analysis, controlling for all sibling shared factors and observed maternal age.

However, it should be noted that a causal interpretation rests on the assumption— common to all sibling designs—that non-shared confounders are adequately controlled. This assumption is unlikely to be true, considering we have not accounted for any non-shared confounders except maternal age. The code to perform the analyses, including estimation of both relative and absolute effects, is provided in the Appendix.

## Discussion

Sibling analysis can be a powerful approach for addressing unmeasured confounding shared within families. In this paper, we have outlined how marginal between-within models can be used to extend sibling analysis to produce clinically meaningful estimates for binary and time-to-event outcomes. By providing both relative measures (e.g., hazard ratios) and absolute measures (e.g., risk differences, number needed to harm), researchers may offer a more complete and interpretable account of their findings. This approach could more appropriately describe the underlying (putative) causal relationship and make it easier for readers to understand the reported associations. While odds ratios and hazard ratios can be informative, exclusive reliance on them may be problematic in many cases.^12^

That said, the interpretation of sibling analysis—like any observational method—depends on strong assumptions^1^. Importantly, it assumes that there are no unmeasured non-shared confounders.^13^ In addition, sibling analyses are susceptible to various forms of bias that can drive estimates both towards or away from the null.^1^ For our applied example of smoking and infant mortality, it seems plausible that carry-over effects could bias the results^14^. However, on a case-by-case basis, it can sometimes be qualitatively argued that an estimate obtained from sibling analysis is less biased than that obtained from a conventional analysis, as it is robust to unobserved confounders shared between siblings.

The marginalized between-within model is less sensitive to some of the generalizability threats to sibling analysis, such as effect modification by shared familial confounders and selection on covariate-discordant families^9^. However, it may still be that the population-level measures estimated from sibling analysis do not directly transport to the entire target population, even if internally valid in the sample contributing to the sibling analysis. For example, if the target population also includes families with fewer than two children, and the causal effect of the exposure on the outcome is materially different in these families, then the sibling analysis estimates may not generalize to the target population. Whether this transportability issue is important should be considered on a case-by-case basis. Finally, it may be noted that between-within models make more assumptions than the conditional logistic regression and stratified Cox regression, in that the latter do not require any assumptions about the functional form or distribution of the baseline. However, this is a necessary assumption if one is to calculate absolute measures.

A challenging aspect of all survival models is the specification of the baseline hazard^15^. In our applied example, we used Breslow’s estimator of the baseline cumulative hazard. Alternative options include assuming a parametric baseline hazard (e.g., a Weibull distribution) or parametrically estimating it using splines (see Appendix)^16–18^. The choice of the baseline hazard should be documented, as it has implications for the estimation of the absolute occurrence of the outcome. Additionally, the method used to compute standard errors can vary by software and modeling choice. Some software offers built-in functions for robust standard errors and delta method recombination (e.g., *stdReg*^*10*^ in R and *standsurv* in Stata), while others may require bootstrapping to account for the sampling variability in the baseline (e.g., *margins* with Breslow’s estimator in Stata). Users may need to be aware of these subtle differences, especially for smaller sample sizes and instances with strong clustering.

## Conclusion

We have presented an overview of sibling analysis models and demonstrated how the marginal between-within framework could be used to estimate absolute measures and clinically meaningful metrics for both binary and time-to-event outcomes. By moving beyond relative measures alone, this approach may enhance the interpretability and practical utility of sibling analyses, allowing researchers to more effectively communicate the magnitude and potential impact of observed associations.

## Data Availability

The data used for the applied example are publicly unavailable according to regula-tions under Swedish law. Readers interested in obtaining microdata may seek similar ap-provals and inquire through Statistics Sweden.

## Statements and Declarations

### Ethics statement

All research was performed in accordance with relevant guidelines/regulations. The applied analysis was approved by the Regional Ethical Review Board in Stockholm (2020-05516).

### Author Contributions

All authors contributed to the conceptualization of the study. VHA performed the analysis, drafted the original manuscript, and curated the applied data. All authors critically revised the manuscript for important intellectual content and contributed to the development of the study methodology. VHA had full access to all the data in the study and takes responsibility for the integrity of the data and the accuracy of the data analysis. All authors read and approved of the final manuscript.

### Availability of data and material

The data used for the applied example are publicly unavailable according to regulations under Swedish law. Readers interested in obtaining microdata may seek similar approvals and inquire through Statistics Sweden.

