## Appendix for "Moving beyond risk ratios in sibling analysis: estimating clinically useful measures from family-based analysis"

### Equations for clinically useful measures

#### Binary outcomes – logistic regression

The average treatment effect comparing exposure X=1 versus X=0, adjusting for observed covariates Z and all shared familial factors, can be expressed as:

$$ATE = E\{P(Y_{ij} = 1 | X_{ij} = 1, Z_{ij}, \bar{X}_i, \bar{Z}_i)\} - E\{P(Y_{ij} = 1 | X_{ij} = 0, Z_{ij}, \bar{X}_i, \bar{Z}_i)\}$$

And:

$$E\{P(Y_{ij} = 1 | X_{ij} = x, Z_{ij}, \bar{X}_i, \bar{Z}_i)\} = \frac{1}{N}\sum_{i=1}^{n}\sum_{j=1}^{n_i}\frac{exp[a + \beta_1(X_{ij} = x) + \theta_1\bar{X}_i + \beta_2 Z_{ij} + \theta_2\bar{Z}_i)]}{1 + exp[a + \beta_1(X_{ij} = x) + \theta_1\bar{X}_i + \beta_2 Z_{ij} + \theta_2\bar{Z}_i)]}$$

Where:

- $i \in \{1, \ldots, n\}$ indexes families (groups)
- $j \in \{1, \ldots, n_i\}$ indexes individuals within family $i$
- $N = \sum_{i=1}^{n} n_i$ is the total number of individuals
- $a$ is the fixed intercept common to all families
- $\beta_1, \beta_2$ capture within-family associations for exposure and covariates
- $\theta_1, \theta_2$ represent between-family effects for exposure and covariates

That is, take the average predicted outcome probability with everyone's treatment value set to 1 and subtract the average predicted outcome probability with everyone's treatment value set to 0. This algorithm corresponds to parametric g-computation, also known as (g-)standardization or the parametric g-formula.

The attributable fraction (also known as the population attributable fraction), representing the proportion of cases that would be avoided if the exposure were eliminated from the population as a whole, is given by:

$$AF = 1 - \frac{E\{P(Y_{ij} = 1 | X_{ij} = 0, Z_{ij}, \bar{X}_i, \bar{Z}_i)\}}{E\{P(Y_{ij} = 1 | X_{ij}, \bar{X}_i, Z_{ij}, \bar{Z}_i)\}}$$

The denominator corresponds to the predicted risk in the population (conditional on covariates), and the numerator reflects the risk if everyone was unexposed (X=0).

The number needed to treat (or harm) for the prospective treatment of the entire population is the inverse of the absolute risk difference:

$$NNT = \frac{1}{E\{P(Y_{ij} = 1 | X_{ij} = 1, Z_{ij}, \bar{X}_i, \bar{Z}_i)\} - E\{P(Y_{ij} = 1 | X_{ij} = 0, Z_{ij}, \bar{X}_i, \bar{Z}_i)\}}$$

#### Time-to-event outcomes – Cox regression

The average treatment effect at time $t$, comparing a hypothetical population where everyone is exposed (X=1) to one where no one is exposed (X=0), after controlling for non-shared confounders $Z_{ij}$ and all factors shared within families, can be expressed as:

$$ATE(t) = E\{S_{ij}(t | X_{ij} = 1, Z_{ij}, \bar{X}_i, \bar{Z}_i)\} - E\{S_{ij}(t | X_{ij} = 0, Z_{ij}, \bar{X}_i, \bar{Z}_i)\}$$

Where, $S_{ij}(t| \cdot)$ denotes the survival for individual $j$ from family $i$, conditional on the specified exposure and non-shared confounders $Z_{ij}$ and all factors shared within families. The average survival can be estimated as:

$$E\{S_{ij}(t | X_{ij} = x, Z_{ij}, \bar{X}_i, \bar{Z}_i)\} = \frac{1}{N}\sum_{i=1}^{n}\sum_{j=1}^{n_i} exp[-\Lambda_0(t) \cdot exp(a + \beta_1(X_{ij} = x) + \theta_1\bar{X}_i + \beta_2 Z_{ij} + \theta_2\bar{Z}_i))]$$

Where:

- $i \in \{1, \dots, n\}$ indexes families (groups)
- $j \in \{1, \dots, n_i\}$ indexes individuals within family $i$
- $N = \sum_{i=1}^{n} n_i$ is the total number of individuals
- $a$ is the fixed intercept common to all families
- $\beta_1, \beta_2$ capture within-family associations for exposure and covariates
- $\theta_1, \theta_2$ represent between-family effects for exposure and covariates
- $\Lambda_0(t) = \int_0^t \lambda_0(u)\, du$ is the cumulative baseline hazard

In other words, $ATE(t)$ represents the expected difference in cumulative survival probabilities at time $t$, had we hypothetically exposed or unexposed the entire population, assuming there are no other confounders.

The attributable fraction at time $t$, reflecting the proportion of events by time $t$ that could be avoided if no one were exposed, is given by:

$$AF(t) = 1 - \frac{E[1 - S_{ij}(t|X_{ij} = 0, Z_{ij}, \bar{X}_i, \bar{Z}_i)]}{E\left[1 - S_{ij}\left(t\middle|X_{ij}, \bar{X}_i, Z_{ij}, \bar{Z}_i\right)\right]}$$

Lastly, the average number of individuals who need to be exposed to prevent (or cause, in the case of harm) one event by time $t$, is as inverse of the survival difference between exposed and unexposed groups by $t$:

$$NNT(t) = \frac{1}{S\left(t\middle|X_{ij} = 1, Z_{ij}, \bar{X}_i, \bar{Z}_i\right) - S(t|X_{ij} = 0, Z_{ij}, \bar{X}_i, \bar{Z}_i)}$$

For harmful exposures, the number needed to harm (NNH) may be calculated by switching the denominator to subtracting the survival under the unexposed scenario from the exposed one.

### Stata code for the marginal between-within analysis

#### Data set-up

To perform all analysis, we have used the following six variables:

| Variable | Description |
|---|---|
| birthdate | Birth date of the child |
| smoking | 0/1 indicating if the mother self-reported smoking at the first antenatal visit |
| matage | Maternal age, zero centered |
| family_id | A unique family identifier (shared among full siblings) |
| death_365 | 0/1 indicating if the child died the first year of life |
| dateexit | Censor date of the child (either death date or end of study date: 31dec2021) |

The first step is to derive the family-level between term for smoking status ($\bar{X}_j$) and maternal age ($\bar{Z}_j$):

```
// Generate between-terms
egen between_smoking = mean(smoking), by(family_id)
egen between_matage = mean(matage), by(family_id)
```

#### Marginal between-within logistic regression

Analyzing mortality as a binary outcome we can implement the marginal between-within logistic model with cluster robust errors.

```
// Fit marginal between-within logistic model
logistic death_365 i.smoking between_smoking matage between_matage, vce(cluster family_id)
```

##### Calculating risk, ATE, AF, and NNT(H) for binary outcomes

From the marginal between-within logistic regression, we can post-estimate the average treatment effect, attributable fraction, and the number needed to treat (number needed to harm, in the case of smoking).

```
// Obtain standardized probabilities across smoking (X) and globally
margins smoking,  grand coeflegend post
// Average treatment effect (difference in probability of event)
lincom _b[1.smoking]-_b[0bn.smoking], cformat(%9.4f)
// Attributable fraction
nlcom 1-(_b[0bn.smoking]/_b[_cons]), cformat(%9.4f)
// Number needed to treat (harm)
nlcom 1/(_b[1.smoking]-_b[0bn.smoking]), cformat(%9.0f)
```

#### Marginal between-within Cox proportional hazards regression

Analyzing mortality as a time-to-event outcome we can implement the marginal between-within Cox model with cluster robust errors:

```
// Declare data to be survival-time data
stset dateexit, failure(death_365== 1) origin(time birthdate  -1)  exit(time birthdate + 365.25)
// Fit marginal between-within Cox regression with clustered standard errors, predicting the baseline survival into variable S0 (note: S0 is predicted when all covariates are set to zero, hence why it may be useful to zero center continous variables – see further description of this in the Stata documentation)
stcox  i.smoking between_smoking matage between_matage, basesurv(S0) vce(cluster family_id)
```

**Calculating 1-Survival, ATE, AF, and NNT(H) for time-to-event outcomes**

From this, we can calculate the absolute outcome functions at different follow-up times. For example, at one year of follow-up (365.25 days):

```
// Find baseline survival when _t=366.25 (i.e., 365.25+1 day to include the first day of life)
summ S0 if _t<=366.25, meanonly
local S0 = r(min)
// Obtain standardized survivals across smoking (X)
margins smoking, expression(1-`S0'^exp(predict(xb)))  grand coeflegend post nose
// Average treatment effect (difference in survivals)
lincom _b[1.smoking]-_b[0bn.smoking], cformat(%9.4f)
// Attributable fraction
nlcom 1-(_b[0bn.smoking]/_b[_cons]), cformat(%9.4f)
// Number needed to treat (harm)
nlcom 1/(_b[1.smoking]-_b[0bn.smoking]), cformat(%9.0f)
```

This procedure can then be repeated by setting different follow-up times (if _t<=28, for example). Note that this code does not consider the sample variability of the baseline survival; in small sample sizes, this could be accounted for by bootstrapping the entire procedure.

### R code for the marginal between-within analysis

#### Data set-up

To perform all analysis, we have used the following six variables, together with the *survival* and *stdReg* packages:

| Variable | Description |
|---|---|
| birthdate | Birth date of the child |
| smoking | 0/1 indicating if the mother self-reported smoking at the first antenatal visit |
| matage | Maternal age, zero centered |
| family_id | A unique family identifier (shared among full siblings) |
| death_365 | 0/1 indicating if the child died the first year of life |
| dateexit | Censor date of the child (either death date or end of study date: 31dec2021) |

The first step is to derive the family-level between term for smoking status ($\bar{X}_j$), and for maternal age ($\bar{Z}_j$):

```
#Generate between-term
data$between_smoking <- ave(data$smoking, data$family_id, FUN=mean)
data$between_matage <- ave(data$matage, data$family_id, FUN=mean)
```

#### Marginal between-within logistic regression

Analyzing mortality as a binary outcome we can implement the marginal between-within logistic model:

```
#Fit marginal between-within logistic model
fit <- glm(formula=death_365~smoking+between_smoking+matage+between_matage, family="bino-
mial", data=data)
```

#### Calculating risk, ATE, AF, and NNT(H) for binary outcomes

We can then post-estimate the average treatment effect, attributable fraction, and the number needed to treat (number needed to harm, in the case of smoking) using the stdReg package by Sjölander[1,2]:

```
#Obtain standardized probabilities across smoking (X)
fit.std <- stdGlm(fit=fit, data=data, X="smoking", x=c(NA,0,1), cluster="family_id")
summary(fit.std)
#Average treatment effect (difference in probability of event)
summary(fit.std, contrast="difference", reference=0)
#Define attributable fraction
AF <- function(est) {
  p <- est[1]
  p0 <- est[2]
  af <- 1-p0/p
  return(af)
```

```
}
#Calculate the attributable fraction
AFest <- AF(fit.std$est)
AFest
#Calculate 95% confidence intervals of the AF
confint(object=fit.std, fun=AF, level=0.95)
#Define NNT (number needed to harm in this case, according to applied example)
NNT <- function(est) {
  p <- est[2]
  p1 <- est[3]
  nnt <- 1/(p1-p)
  return(nnt)
}
#Calculate NNT
NNTest <- NNT(fit.std$est)
NNTest
#Calculate 95% confidence intervals of the NNT
confint(object=fit.std, fun=NNT, level=0.95)
```

##### Marginal between-within Cox proportional hazards regression

Analyzing mortality as a time-to-event outcome we can implement the marginal between-within flexible parametric model with cluster robust errors:

```
#Create a survival time indicator
data$surv_time <- pmin(as.numeric(data$dateexit - data$birthdate), 365.25)
#Fit marginal between-within Cox regression with clustered standard errors
fit <- coxph(formula=Surv(surv_time, death_365)~smoking+between_smoking+matage+be-
tween_matage, data=data, ties=c("breslow"), cluster=family_id)
```

##### Calculating 1-Survival, ATE, AF, and NNT(H) for time-to-event outcomes

From this, we can calculate the absolute outcome functions at different follow-up times. For example, at one year of follow-up (365.25 days):

```
#Perform standardization at _t=365.25
fit.std <- stdCoxph(fit=fit, data=data, X="smoking", x=c(NA,0,1), t=365.25)
summary(fit.std, contrast="difference", reference=1)
#Define AF
AF <- function(est) {
p <- 1-est[, 1]
p0 <- 1-est[, 2]
af <- 1-p0/p
return(af)
}
#Calculate AF
AFest <- AF(fit.std$est)
AFest
#Calculate 95% confidence intervals of the AF
confint(object=fit.std, fun=AF, level=0.95)
```

```
#Define the number needed to treat (harm in this case)
NNT <- function(est){
  p <- 1-est[, 2]
  p1 <- 1-est[, 3]
  nnt <- 1/(p1-p)
  return(nnt)
}
#Calculate the number needed to treat (harm in this case)
NNTest <- NNT(fit.std$est)
NNTest
#Calculate 95% confidence intervals of the NNT
confint(object=fit.std, fun=NNT, level=0.95)
```

#### Marginal between-within flexible parametric regression

Beyond Breslow's estimator, the baseline function can be estimated parametrically using restricted cubic splines. In this approach, the splines allow for flexible curvature but are constrained to be linear before the first knot and after the last.[3] The standard (non-clustered) flexible parametric model for the log cumulative hazard can be expressed as:

$$ln(\Lambda_0[t|X,Z]) \ = ln(\Lambda_0(t)|\gamma, k_0) + \beta_1 X + \beta_2 Z$$

Here, $\Lambda_0(t)$ is the baseline cumulative hazard, modeled using restricted cubic splines with $k_0$ knots. For example, with 4 knots:

$$ln(\Lambda_0(t)|\gamma, k_0) \ = \gamma_0 + \gamma_1 s_1 + \gamma_2 s_2 + \gamma_3 s_3$$

where $s_1, s_2, s_3$ are the spline basis functions, and the $\gamma$ coefficients are estimated from the data, and $k_0$ a vector of knot positions. It is important to note that, as currently specified, this is a proportional hazard model[4], and the estimated hazard ratios will closely approximate those obtained via Cox regression. The integration of the flexible parametric model into a marginal between-within framework is straightforward and leads to the following specification:

$$log\left(\Lambda_{ij}\left[t|X_{ij}, Z_{ij}, \bar{X}_i, \bar{Z}_i\right]\right) \ = log(\Lambda_0(t)|\gamma, k_0) \ + \beta_1 X_{ij} + \theta_1 \bar{X}_i + \beta_2 Z_{ij} + \theta_2 \bar{Z}_i$$

In this model, $log\left(\Lambda_0(t)\right)$ represents the shared (across all families) log cumulative baseline hazard; $\beta_1$ captures the within-family association between the exposure and the outcome, adjusting for the measured non-shared confounder $Z$ and for shared familial factors.

It is important to note that this model differs from the conditional frailty model proposed by Dahlqwist et al. [5] in that it does not rely on a specified form for the shared frailty distribution (e.g., Weibull-gamma). Moreover, the marginal between-within flexible parametric model is typically more computationally efficient, as it avoids the intensive integration required for fitting frailty models. The downside of this model is that it requires a parametric estimation of the baseline hazard function and that one must specify an appropriate number of knots and the positions of such knots, albeit some effect estimates (e.g., hazard ratios) may be insensitive to the number of knots[6,7].

##### Stata code for the marginal between-within flexible parametric regression

We can implement the marginal between-within flexible parametric model in Stata with cluster robust errors using Lambert's Stata package *stpm2*[8] and *standsurv*:

```
// Declare data to be survival-time data
stset dateexit, failure(death_365== 1) origin(time birthdate  -1)  exit(time birthdate + 365.25)
// Fit flexible marginal between-within model with four degrees of freedom.
stpm2 smoking between_smoking, df(4) scale(hazard) eform  vce(cluster family_id)
```

From this, we can calculate standardized survival functions across follow-up time.

```
// Calculate standardized survival functions across follow-up
range time 0 365 366 /// Generate time variable over which to predict
standsurv, timevar(time) failure ci /// request failure function (1-s[t]) over time together with 95% CI
                                        at1(smoking 0) /// The first function is when everyone is set to smoking=0
                                        at2(smoking 1) /// The second function is when everyone is set to smoking=1
                          contrast(difference) /// Calculate the contrast (the average treatment effect)
                                                      atvar(std_smoking0 std_smoking1) // store results
// Show results at 7, 28, 180, and 365 days of follow-up
list time *smoking0* *smoking1* *contrast* if inlist(time,7, 28, 180, 365)
```

Unlike the Stata code for the marginal between-within Cox regression, the above uses the delta method to calculate the empirical standard errors, or could be implemented with m-estimation, so bootstrapping is not required for valid inferences.
